# Detection of Suicidality Through Privacy-Preserving Large Language Models

**DOI:** 10.1101/2024.03.06.24303763

**Authors:** Isabella Catharina Wiest, Falk Gerrik Verhees, Dyke Ferber, Jiefu Zhu, Michael Bauer, Ute Lewitzka, Andrea Pfennig, Pavol Mikolas, Jakob Nikolas Kather

## Abstract

**Importance:** Attempts to use Artificial Intelligence (AI) in psychiatric disorders show moderate success, high-lighting the potential of incorporating information from clinical assessments to improve the models. The study focuses on using Large Language Models (LLMs) to manage unstructured medical text, particularly for suicide risk detection in psychiatric care.

**Objective:** The study aims to extract information about suicidality status from the admission notes of electronic health records (EHR) using privacy-sensitive, locally hosted LLMs, specifically evaluating the efficacy of Llama-2 models.

**Main Outcomes and Measures:** The study compares the performance of several variants of the open source LLM Llama-2 in extracting suicidality status from psychiatric reports against a ground truth defined by human experts, assessing accuracy, sensitivity, specificity, and F1 score across different prompting strategies.

**Results:** A German fine-tuned Llama-2 model showed the highest accuracy (87.5%), sensitivity (83%) and specificity (91.8%) in identifying suicidality, with significant improvements in sensitivity and specificity across various prompt designs.

**Conclusions and Relevance:** The study demonstrates the capability of LLMs, particularly Llama-2, in accurately extracting the information on suicidality from psychiatric records while preserving data-privacy. This suggests their application in surveillance systems for psychiatric emergencies and improving the clinical management of suicidality by improving systematic quality control and research.

**Key Points:** *Question:* Can large language models (LLMs) accurately extract information on suicidality from electronic health records (EHR)?

*Findings:* In this analysis of 100 psychiatric admission notes using Llama-2 models, the German fine-tuned model (Emgerman) demonstrated the highest accuracy (87.5%), sensitivity (83%) and specificity (91.8%) in identifying suicidality, indicating the models’ effectiveness in on-site processing of clinical documentation for suicide risk detection.

*Meaning:* The study highlights the effectiveness of LLMs, particularly Llama-2, in accurately extracting the information on suicidality from psychiatric records, while preserving data privacy. It recommends further evaluating these models to integrate them into clinical management systems to improve detection of psychiatric emergencies and enhance systematic quality control and research in mental health care.

## Introduction

Attempts to apply artificial intelligence (AI) and machine learning to psychiatric disorders have yielded moderate accuracies due to small effect sizes and high heterogeneity.^1^ Nevertheless, improving prediction models by incorporating clinical assessments seems to enable clinical applications.^2^ However, a significant challenge arises from the nature of clinical data: Medical free text, especially in psychiatry, encapsulates a wealth of information about a patient’s pathology and well-being by unveiling its structure of thinking and feeling. This information is vital but often remains inaccessible for scalable analysis due to its unstructured nature. The inability to effectively analyze this text on a large scale potentially leads to missed opportunities in clinical decision making and research.

Recent studies have emphasized the significant impact of advanced technology on managing unstructured medical data^3^. Specifically, the use of large language models (LLMs) has garnered significant attention.^4^ Unlike previously used methods of natural language processing that require decomposing the text and substantial feature engineering,^5^ LLMs are AI models primarily designed to understand and generate text.^6^ They are trained on vast amounts of text data, allowing them to learn the statistical patterns and relationships within language.^7^

Accounting for nearly half of all emergency psychiatric admissions,^8^ suicide is one of the most tragic complications of psychiatric care and is often preventable. Sustained efforts can lead to major reductions in in-patient suicides, from 4.2 to 0.74 per 100,000 admissions.^9^ Here, we hypothesize that automated tools could help identify in-patient suicide risk using underexploited clinical records. Moreover, beyond clinical application, LLM might automatically identify and extract suicidality from EHR to enhance research.

## Methods

We systematically extracted n=100 randomly selected text-based admission notes of inpatients treated in and discharged from the acute psychiatric ward of the Department of Psychiatry and Psychotherapy at the University Hospital Carl Gustav Carus Dresden between 1 January and 31 December 2023, representing 54 female and 46 male patients with an average age of 50 years (standard deviation 23.8 years) ranging from 18 to 96 years of age. The most prevalent ICD-10 main diagnoses were major depressive disorder (21%), psychotic disorders (20%) and dementia (17%), borderline personality disorder (9%), schizoaffective disorder (8%), alcohol use disorder (8%) and others (17%). We ensured data privacy by installing Llama-2 via the llama.cpp framework on a local hospital computer. We extracted the suicidality status from psychiatric admission notes using three different Llama-2-based models: the standard Llama-2 70b chat model adapted to allow deployment on low-resource consumer hardware,^10^ as well as two versions of Llama-2 that were specifically fine-tuned for the German language (“Sauerkraut”^11^ and “Emgerman”^12^). We compared the models’ results to a ground truth consensus which was established by a resident (FGV) and a consultant psychiatrist (PM) as a binary variable (suicidal / not suicidal). Suicidality was defined as either suicidal thoughts, ideation, plans or attempt by admission.

We applied a step-by-step approach to prompt engineering, as prompt engineering can substantially improve the performance of LLMs.^13^ The first prompt simply asked about suicidality in reports (P0). In the second prompt, we added fictional examples and explanations. We started with one example (P1) and added one example (P2) at a time with three examples as a maximum (P3). (See prompts in **Supplementary Table 1**). After achieving improved performance, we incorporated a chain-of-thought approach, where the model processes its own output one more time, for P3 (P4). To obtain reliable estimates, we used bootstrapping, a statistical resampling technique, with 10.000 iterations.

All research procedures were conducted in accordance with the Declaration of Helsinki. Ethics approval was granted by the ethics committee of Technical University Dresden, reference number BO-EK-400092023. All source codes are available at https://github.com/I2C9W/LLM4Psych/tree/v0.1.0.

## Results

Llama-2 extracted suicidality from psychiatric reports with high accuracy across all five prompt designs and all three models tested. The highest overall accuracy was achieved by one of the german fine-tuned Llama-2 models (“Emgerman”), which correctly identified suicidality status in 87.5% of the reports. With a sensitivity of 83% and a specificity of 92%, it demonstrated the highest balanced accuracy of all models (87.4%) (**Figure 2A**).

**Figure 1.**
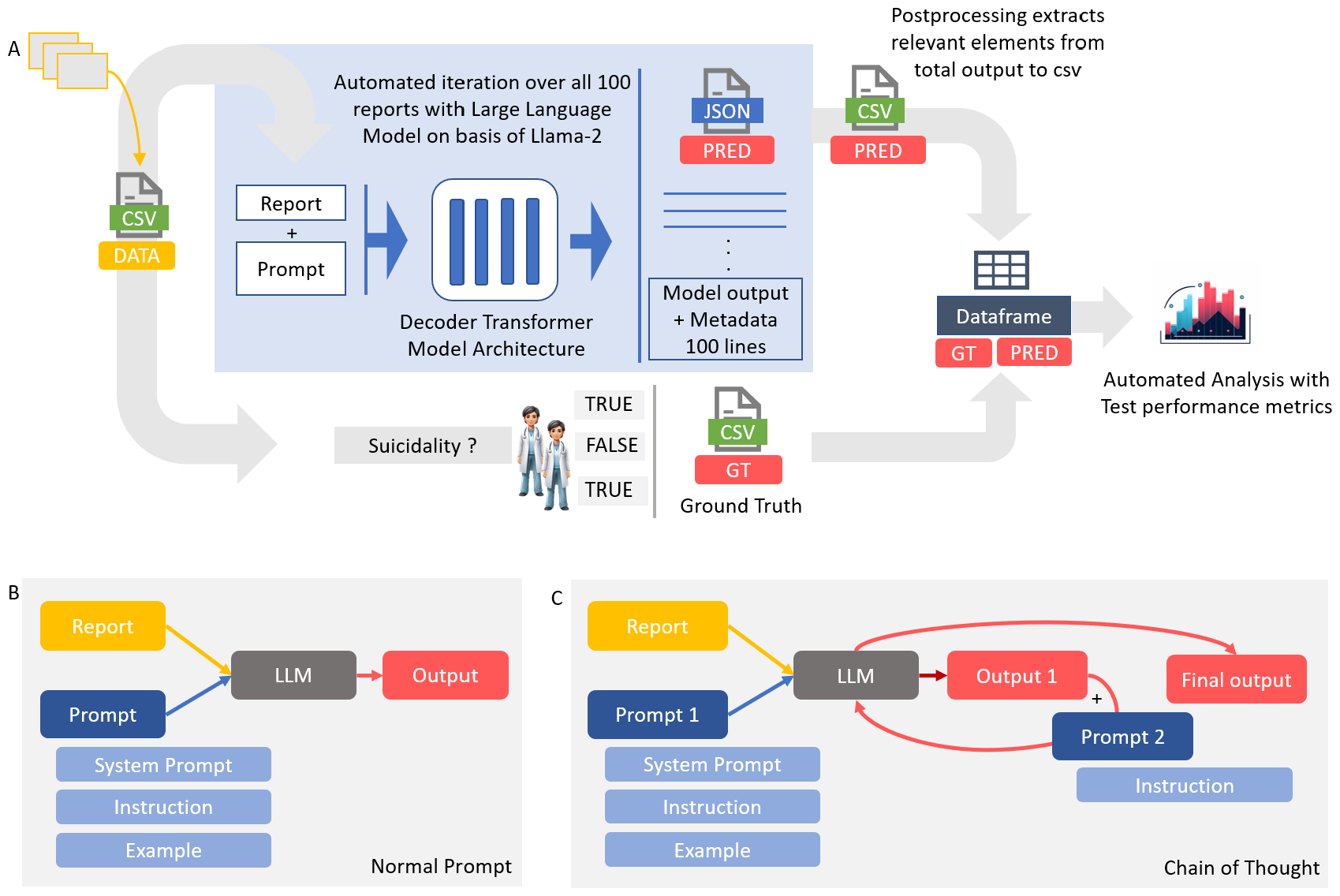
Experimental Setup. **A** displays the information extraction pipeline. The psychiatry reports (n=100) were transferred to a csv table. Our pipeline then iterates over all reports with the predefined prompt and outputs a JavaScript Object Notation-File (JSON) file with all Large Language Model (LLM) outputs (PRED). The relevant classes (suicidality present: yes or no) were then extracted from the LLM output, which was more verbose in some cases. These outputs were then transferred to a pandas dataframe and automatically compared to the expert-based ground truth (GT). **B** depicts the initial prompting strategy. One prompt and one report were given to the model at the same time. Every prompt contained a system prompt with general instructions and a specific question to the report (Instruction) **C** shows the chain of thought approach: The psychiatry report with our prompt was fed into the LLM, which generated a first output. With a second prompt and a predefined answering grammar, the model was fed its own output and again forced to generate a certain, json based output structure. This final output then underwent performance analysis. Icon Source: Midjourney.

**Figure 2.**
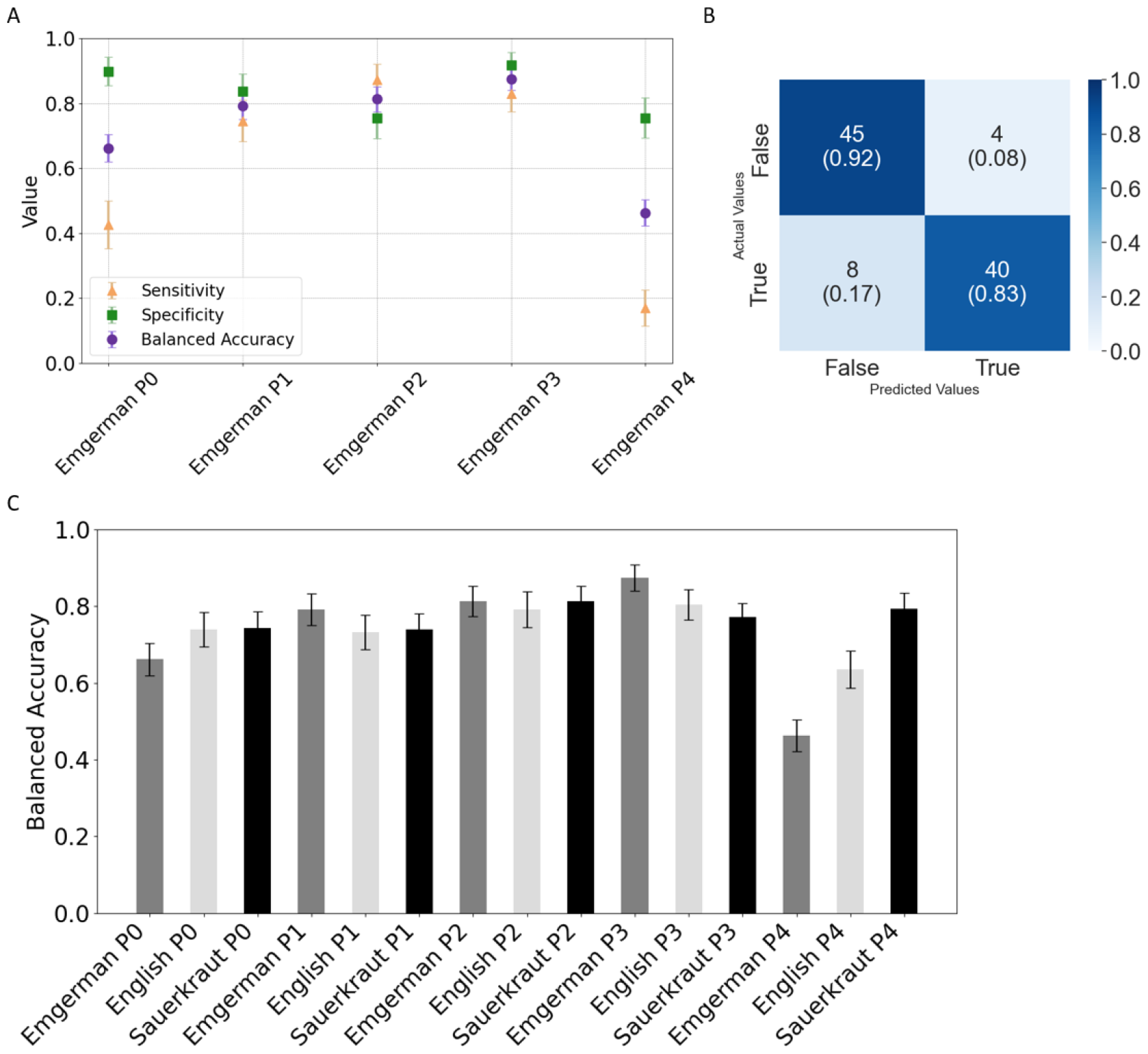
Performance of german-language fine-tuned Llama-2 model. **A** depicts Sensitivity, Specificity and balanced Accuracy score for five different prompting strategies. With P0, the model was simply asked to provide the answer if suicidality was present from the report, P1, P2 and P3 provided one, two or three examples to the model. P4 applied a chain-of-thought approach, where the model was asked twice, with the first model output as input for the second run. **B** The confusion matrix represents the performance of the LLM indicating the presence of suicidality based on the examined admission notes (n=100) with a sensitivity of 83% as well as specificity of 92% for P3, a prompt that included three examples. **C** The bar chart shows the balanced accuracies for all models and prompt engineering attempts. Error bars show the 95% confidence interval of the bootstrapped samples.

The confusion matrix (**Figure 2B**) also highlights areas for model improvement, particularly in reducing false negatives.

To improve the performance, we designed the prompts and developed five different prompting strategies that were tested for all three models (**Figure 2C**). The simplest prompt, which contained only a “system prompt” framing the model in its role (“You are an attentive medical assistant with specialized knowledge in psychiatry (…)”, one report at a time and the ultimate question of interest (“Is the patient suicidal? Answer yes or no. (…)”), yielded the highest sensitivity in the German fine-tuned Llama-2 model “Sauerkraut” (sensitivity: 87.5%, specificity: 61.2%, balanced accuracy: 74.4%). It was immediately followed by the standard English Llama-2 chat model, with a sensitivity of 85.1%, specificity of 63% and a balanced accuracy of 74.1%. The Emgerman model had a worse sensitivity of 42.6%, but the highest specificity of 98.8%. Not all models improved when examples were added to the prompt, allowing for in-context-learning. The Emgerman model improved substantially by adding more examples, with the lowest balanced accuracy in the prompt with no examples (66.2%) and the highest balanced accuracy in the prompt with three examples given (87.4%). The English model was robust, showing similar balanced accuracies for prompts with none, one, two or three examples (P0: 74.1%, P1: 73.3%, P2: 79.3%, P3: 80.3%). The “Sauerkraut” model improved with adding examples but achieved its maximum performance with two examples in the prompt. The use of the chain-of-thought approach did not improve performance (Sensitivities: “Emgerman” P4 17%, “English” P4 63.8%, “Sauerkraut” P4 80.9%. Specificities: “Emgerman” P4 75.5%, “English” P4 63.3%, “Sauerkraut” P4 77.6%. (**Table 1)**). In fact, all models deteriorated, except for the “Sauerkraut” model, which was not affected negatively by this approach.

**Table 1.** Performance Metrics of all three tested models (“Emgerman”, “Sauerkraut”, “English”) with the five prompt variations (P0-P4). All results have been obtained by 10.000 fold bootstrapping, therefore means and standard deviations are given.

| Model | Accuracy Mean | Accuracy Std | PPV Mean | PPV Std | Sensitivity Mean | Sensitivity Std | Specificity Mean | Specificity Std | NPV Mean | NPV Std | F1 Score Mean | F1 Score Std | Balanced Accuracy Mean | Balanced Accuracy Std |
| --- | --- | --- | --- | --- | --- | --- | --- | --- | --- | --- | --- | --- | --- | --- |
| Emgerman P0 | 0.667 | 0.048 | 0.8 | 0.082 | 0.426 | 0.073 | 0.898 | 0.044 | 0.62 | 0.057 | 0.552 | 0.072 | 0.662 | 0.042 |
| Emgerman P1 | 0.793 | 0.041 | 0.815 | 0.059 | 0.746 | 0.064 | 0.837 | 0.053 | 0.775 | 0.058 | 0.777 | 0.049 | 0.792 | 0.041 |
| Emgerman P2 | 0.812 | 0.04 | 0.773 | 0.058 | 0.872 | 0.049 | 0.754 | 0.062 | 0.86 | 0.053 | 0.818 | 0.042 | 0.813 | 0.039 |
| Emgerman P3 | 0.875 | 0.034 | 0.907 | 0.044 | 0.83 | 0.055 | 0.918 | 0.039 | 0.849 | 0.049 | 0.865 | 0.039 | 0.874 | 0.034 |
| Emgerman P4 | 0.468 | 0.051 | 0.4 | 0.112 | 0.17 | 0.055 | 0.755 | 0.062 | 0.486 | 0.057 | 0.236 | 0.069 | 0.463 | 0.041 |
| English P0 | 0.741 | 0.046 | 0.7 | 0.061 | 0.851 | 0.052 | 0.629 | 0.072 | 0.805 | 0.067 | 0.767 | 0.047 | 0.74 | 0.045 |
| English P1 | 0.731 | 0.045 | 0.703 | 0.062 | 0.792 | 0.059 | 0.672 | 0.067 | 0.767 | 0.066 | 0.743 | 0.049 | 0.732 | 0.045 |
| English P2 | 0.788 | 0.048 | 0.731 | 0.069 | 0.881 | 0.055 | 0.703 | 0.074 | 0.866 | 0.062 | 0.797 | 0.052 | 0.792 | 0.046 |
| English P3 | 0.805 | 0.04 | 0.854 | 0.055 | 0.73 | 0.065 | 0.878 | 0.047 | 0.768 | 0.057 | 0.785 | 0.048 | 0.804 | 0.04 |
| English P4 | 0.635 | 0.049 | 0.625 | 0.07 | 0.638 | 0.071 | 0.633 | 0.068 | 0.646 | 0.069 | 0.629 | 0.058 | 0.636 | 0.049 |
| Sauerkraut P0 | 0.742 | 0.044 | 0.689 | 0.059 | 0.875 | 0.048 | 0.612 | 0.07 | 0.833 | 0.063 | 0.769 | 0.045 | 0.743 | 0.042 |
| Sauerkraut P1 | 0.742 | 0.044 | 0.897 | 0.057 | 0.542 | 0.072 | 0.939 | 0.034 | 0.677 | 0.056 | 0.672 | 0.062 | 0.74 | 0.04 |
| Sauerkraut P2 | 0.815 | 0.039 | 0.858 | 0.054 | 0.749 | 0.062 | 0.878 | 0.047 | 0.781 | 0.056 | 0.798 | 0.047 | 0.814 | 0.039 |
| Sauerkraut P3 | 0.773 | 0.042 | 0.964 | 0.035 | 0.562 | 0.071 | 0.98 | 0.02 | 0.696 | 0.055 | 0.708 | 0.06 | 0.771 | 0.037 |
| Sauerkraut P4 | 0.793 | 0.042 | 0.777 | 0.059 | 0.81 | 0.057 | 0.776 | 0.06 | 0.81 | 0.057 | 0.791 | 0.046 | 0.793 | 0.042 |

PPV=Positive Predictive Value, NPV = Negative Predictive Value, Std= Standard Deviation

## Discussion

We show that LLMs demonstrate remarkable efficacy in identifying and extracting references to suicidality from psychiatric reports. Its performance, in terms of both sensitivity and specificity, was notable and improved progressively with the number of examples provided in the prompt. These findings suggest a significant advancement in the field, highlighting the potential of LLMs to revolutionize the way psychiatric medical text is analyzed. The real-life clinical data taken from an acute care ward in a supra-maximum care facility in a German urban center was processed at the “edge” - with no need for upload to commercial servers or a data-processing cloud - by an open-source model on local servers. This enables a privacy-sensitive data protection strategy in a closed loop, that alleviates concerns about data leaving the care provider’s control.

The good performance levels (**Figure 2**) even in a (medical) domain in which the LLM was not fine-tuned, suggest even greater opportunities with further optimization for mental health, e.g. in dealing with physician-level linguistic idiosyncrasies or abbreviations.^14^ For a clinical application such as suicide risk detection, where false negatives are likely to lead to detrimental outcomes, sensitivity should approach 100%, even at the cost of detecting more false positives. The final risk assessment remains in the judgment of the experienced clinician and further research needs to elucidate risks and challenges. On the other hand, in the case of data extraction for research purposes, correctly identifying 80% of cases (i.e. classification accuracy of 80%) might be adequate to capture a representative cohort. In comparison, randomized clinical trials of major depression may include only 22% of cases from real-life clinical cohorts that meet the eligibility criteria.^15^

Suicide risk was considered a binary parameter. Future research should concentrate on a more detailed outcome that differentiates between overall suicide risk and acute high risk.^16^ Additionally, studies should apply extensive ground truth labeling,^17^ include open cases that have not been proofread and evaluate more comprehensive prompt engineering strategies. However, our results suggest that, at least in the case of Llama-2, more complex prompting with a chain-of-thought approach might degrade performance. For some tasks, simple example prompting that requires very few computing resources may be more suitable. Nevertheless, reproducibility should be tested on a larger external validation sample. Although privacy concerns have been addressed, it is important to note that every LLM approach inherits ethical issues related to bias, trust, authorship, and equitability.^18^ Expert guidelines for development of LLMs for medical purposes should be carefully considered.^19^

## Conclusion

We provide a proof-of-concept analysis for automated extraction of in-patient suicidality from EHR using LLM. Possible applications include early warning and surveillance tools for psychiatric emergencies, preventing information transfer failures, quality assurance and last but not least evaluation of psychiatric symptoms on large clinical “real-world” samples.

## Data Availability

All source codes are available at https://github.com/I2C9W/LLM4Psych/tree/v0.1.0. All data data used in the study cannot be made available because of the sensitive nature of psychiatric medical admission notes.

## Supplement

**Supplementary Table 1.** P0 shows the simplest prompt without examples, P3 is the extended version with three examples in the text to ensure in-context learning. P1 and P2 have one and two examples, respectively. The original German prompt has been translated into English.

| Prompt Abbreviation | Prompt (Translation) |
| --- | --- |
| P0 | "You are an attentive medical assistant with specialized expertise in psychiatry. Below you will find a psychiatric history. Please answer the question.<br>Medical history: {}<br>Question: Is the patient suicidal? Answer yes or no. Give an excerpt from the medical history to justify your answer." |
| P3 | "You are an attentive medical assistant with specialized knowledge in psychiatry. Below you will find a psychiatric history. Please answer the question. First you will be given an example.<br>Example 1: Patient is awake, conscious, friendly and demonstrating a willingness to engage in a conversation. Disorientation of time (-1 day). Oriented to the location, situation and personal identity. Perception, concentration and memory are intact. No evidence of formal thought disorder or abnormal thought content. Mood stable, affect appropriate to the situation and within normal fluctuations. No fears or compulsions. No abnormalities of activity. No feelings of weariness with life or suicidal thoughts. No immediate risk to self or others.<br>Question: Answer yes or no. Give an excerpt from the medical history to justify your answer.<br>ASSISTANT: no. Rationale: Medical history indicates that the patient is not experiencing any feelings of weariness with life or suicidal thoughts. In addition, there is no immediate risk to self or others. In summary, there is no suicidal ideation.<br>Example 2: Awake, conscious. Signs of impaired comprehension and attention. Thought process of intact rate, perseverating and fixated to the precipitating situation and desire to self-discharge. No signs of delusions. Perceptual disorders denied. Mood clearly dysphoric and depressed, without evident mood swings. Psychomotor agitation. No immediate indications of danger to others, acute danger to self in case of suicidal tendencies.<br>Question: Answer yes or no. Give an excerpt from the medical history to justify your answer.<br>ASSISTANT: yes. Rationale: This can be inferred from the last sentence of the medical history: "acute danger to self in case of suicidal tendencies"<br>Example 3: Patient awake, conscious, fully oriented across all 4 dimensions, friendly, presenting himself as willing to provide information, however, mood barely explorable due to drug intoxication, displaying intermittently parathymic and inappropriate affect, laughing out loud during the conversation. Formal and content-related thought process clearly incoherent, signs of hallucinations (looks around the room), no evidence of specific fears or compulsions. Currently clearly not displaying acute suicidal tendencies. Signs of acute danger to others. A risk of harm to both themselves and others in the context of psychotic misinterpretation of reality.<br>Question: Answer yes or no. Give an excerpt from the medical history to justify your answer.<br>ASSISTANT: no. Reason: No, the patient is currently clearly not displaying acute suicidal tendencies<br>Medical history: {}<br>Question: Is the patient suicidal? Answer yes or no. Give an excerpt from the medical history to justify your answer." |

